# Associations with spontaneous and indicated preterm birth in a densely phenotyped EHR cohort

**DOI:** 10.1101/2023.11.29.23299216

**Authors:** Jean M. Costello, Hannah Takasuka, Jacquelyn Roger, Ophelia Yin, Alice Tang, Tomiko Oskotsky, Marina Sirota, John A. Capra

## Abstract

**Background:** Preterm birth (PTB) is the leading cause of infant mortality and follows multiple biological pathways, many of which are poorly understood. Some PTBs result from medically indicated labor following complications from hypertension and/or diabetes, while many others are spontaneous with unknown causes. Previously, investigation of potential risk factors has been limited by lack of data on maternal medical history and the difficulty of classifying PTBs as indicated or spontaneous. Here, we leverage electronic health record (EHR) data (patient health information including demographics, diagnoses, and medications) and a supplemental curated pregnancy database to overcome these limitations. Novel associations may provide new insight into the pathophysiology of PTB as well as help identify individuals who would be at risk of PTB.

**Methods:** We quantified associations between maternal diagnoses and preterm birth using logistic regression controlling for maternal age and socioeconomic factors within a University of California, San Francisco (UCSF), EHR cohort with 10,643 births (*nterm* = 9692, *nspontaneous_preterm* = 449, *nindicated_preterm* = 418) and maternal pre-conception diagnosis phenotypes derived from International Classification of Diseases (ICD) 9 and 10 codes.

**Results:** Eighteen conditions significantly and robustly (False Discovery Rate (FDR)<0.05) associated with PTBs compared to term. We discovered known (hypertension, diabetes, and chronic kidney disease) and less established (blood, cardiac, gynecological, and liver conditions) associations. Type 1 diabetes was the most significant overall association (adjusted p = 1.6×10^-14^, adjusted OR = 7 (95% CI 5, 12)), and the odds ratios for the significant phenotypes ranged from 3 to 13. We further carried out analysis stratified by spontaneous vs. indicated PTB. No phenotypes significantly associated with spontaneous PTB; however, the results for indicated PTB largely recapitulated the phenotype associations with all PTBs.

**Conclusions:** Our study underscores the limitations of approaches that combine indicated and spontaneous births together. When combined, significant associations were almost entirely driven by indicated PTBs, although our spontaneous and indicated groups were of a similar size. Investigating the spontaneous population has the potential to reveal new pathways and understanding of the heterogeneity of PTB.

## 1 Background

Preterm birth (PTB) is the leading cause of infant mortality worldwide (1) and can result in serious acute and long-term health consequences (2,3). There are multiple proposed pathways for preterm birth, but its etiology remains poorly understood (4–7). About two thirds of PTBs in the US are classified as spontaneous preterm while the remaining third are medically indicated (iatrogenic) preterm (8). An indicated preterm birth is typically initiated based on a list of risk factors, which includes preeclampsia, diabetes complications, intrauterine abnormalities, and placental abnormalities (9). Some of these risk factors, such as poorly managed hypertension, may be present prior to pregnancy. Spontaneous preterm birth, by contrast, lacks a defined set of known risk factors, and the pathophysiology behind it remains poorly understood (8).

Maternal risk factors for indicated preterm birth include older maternal age, heart disease, hypertension, diabetes, tobacco use, previous preterm delivery, and socioeconomic factors (8,10). Zheng et al. studied lifestyle factors, obstetric and fetal complications, maternal diseases, and socioeconomic factor associations with preterm birth in 3,147 cases and controls across 15 Chinese hospitals (11). They measured multiple pregnancies, hypertensive disorders, and obstetric disorders to be the strongest predictors of iatrogenic preterm birth, with socioeconomic risk factors such as maternal education and prenatal care access also significant.

Several maternal risk factors for spontaneous preterm birth have been proposed, including prior spontaneous preterm birth, gynecological anatomy variation, short inter-pregnancy interval, and multiple gestations (12). Prior spontaneous preterm birth is the strongest known risk factor. In the United States, racism is a risk factor for spontaneous preterm birth (13), with higher rates among non-Hispanic Black birthing people when compared to white birthing people, including after adjustment for socioeconomic variables (14). Some studies have explored whether gene-gene and/or gene-environment interactions might exist to explain racial disparities, but these studies are limited to cohorts of a few hundred patients (12).

Improved understanding of pathways and clinical factors leading to preterm birth could lead to better interventions to prevent preterm birth, especially spontaneous preterm birth. Investigating pre-pregnancy conditions associated with subsequent PTB has the potential to generate hypotheses about pathways towards PTB. Many large studies of conditions associated with PTB rely on registry data, which provides limited phenotypic information.

EHR databases provide dense phenotyping including demographics, diagnoses, and medications over time that can provide insights difficult to obtain from other data sources. However, EHR systems may not distinguish between spontaneous and indicated deliveries. Nonetheless, EHR data are particularly well-suited to the study of pregnancy (15). For instance, machine learning models have used EHR data to accurately predict preterm birth on thousands of patients (16). While complex machine learning models have great potential to improve obstetric and gynecological care, novel insights from straightforward methods applied to EHR data could more easily translate to pathway discovery and evidence-based care.

In this study, we use logistic regression to measure associations between maternal pre-conception diagnoses and different types of preterm birth using University of California, San Francisco EHR data supplemented with a physician-curated database of delivery information. With this approach, we reproduce widely known preterm birth risk factors including older maternal age and major chronic diseases. Moreover, we discover several new preterm birth associations with less-studied conditions such as decreased white blood count. We also demonstrate that all significant associations with PTB are driven by indicated PTBs and that no diagnoses significantly associate with spontaneous preterm birth.

## 2 Results

### A densely phenotyped preterm birth cohort linked to electronic health records

To identify potential clinical risk factors for PTB, we defined cohorts of preterm and term deliveries based on curated data from the UCSF Perinatal Database (PDB) and linked these to phenotypes from the UCSF electronic health record (EHR) database.

The cohort consisted of 10,643 deliveries to 9,399 individuals from 2001 to 2022 (**Figure 1a**). There were 975 PTBs in the cohort, which we further classified as spontaneous PTBs (n=449) or indicated PTBs (n=418). The remaining 108 PTBs could not be classified. Each of the preterm groups (spontaneous, indicated, all) was compared to term “controls” born at 37 weeks or later (n=9671). More details about the cohorts are provided in the Methods section.

**Figure 1:**
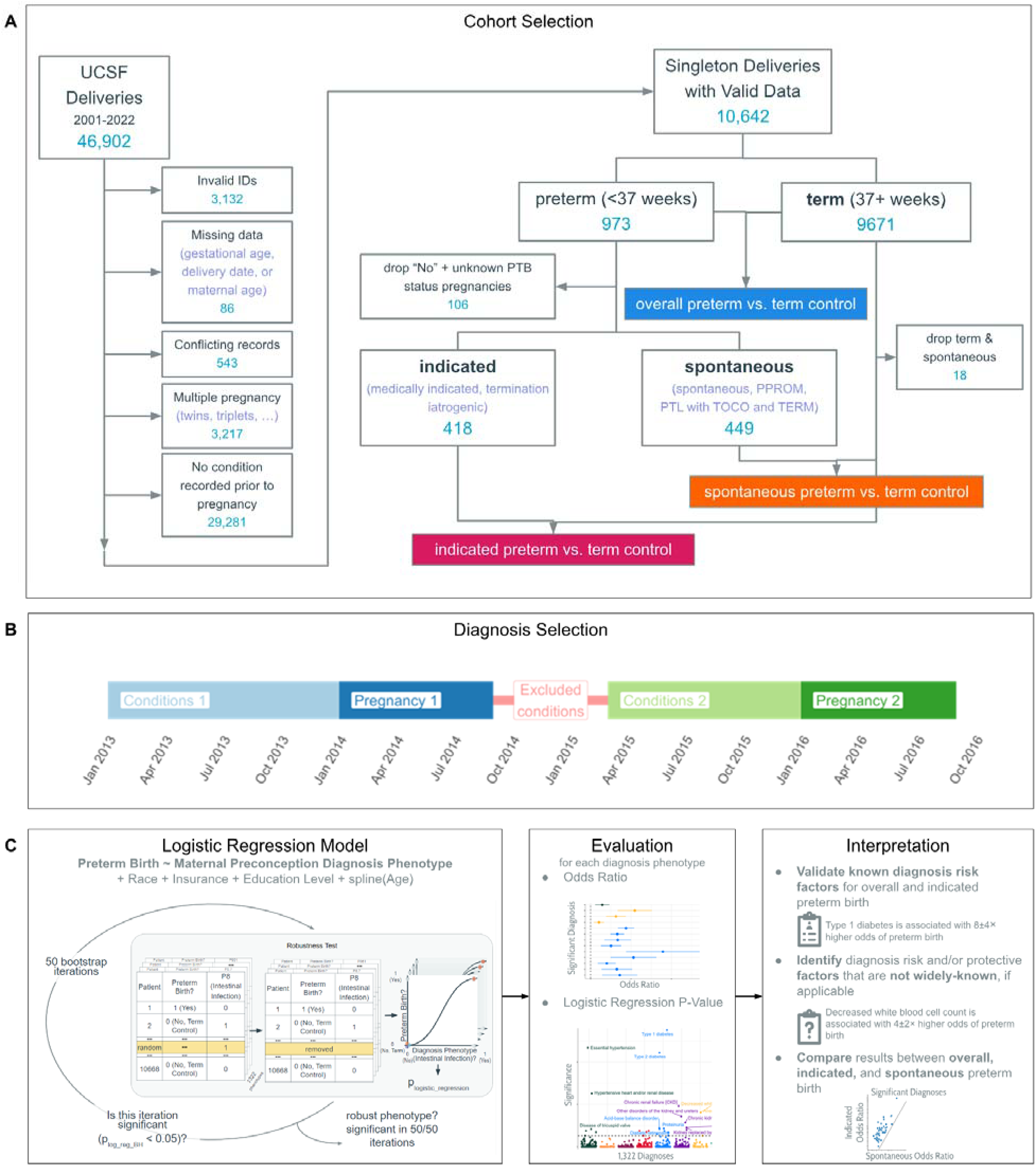
Schematic of the Approach for Testing Associations Between Preterm Birth and Diverse Phenotypes. (A) Criteria for identifying the 10,642 individuals studied and assigning them to overall preterm, indicated preterm, spontaneous preterm, and term groups for subsequent logistic regression analyses. (B) Diagnoses before conception are used in this study. For a person’s first birth (or only birth), diagnoses are recorded from the start of their record until the start of conception. If multiple births are recorded for the same individual, diagnoses for subsequent births are recorded starting 6 months from after the previous delivery to the start of the next conception. (C) Overview of the logistic regression analysis, covariates, evaluation, and interpretation for associations between preterm birth and the 1322 diagnosis phenotypes considered. P-values were adjusted for multiple hypothesis testing and a permutation test was used to ensure associations were robust.

The demographics of the cohort reflected the population of the San Francisco Bay area served by UCSF. Most individuals had more than 12 years of education (84%). A large majority also used private insurance for the delivery (93.2%). The mean maternal age was 34.4 years, and maternal age ranged from 14 years to 55 years. There were no significant differences in maternal age between indicated, spontaneous, and term individuals (**Figure S2a**; pindicated-term = 0.2, pspontaneous-term = 0.1, pindicated-spontaneous = 0.9, Mann-Whitney U test). The two most represented self-reported racial categories were single-race white (48.3%) and single-race Asian/Pacific Islander (25%) (Table 1).

**Table 1:** Demographics. Race, maternal age, maternal education level, insurance status (private, public, or unknown), and diagnosis distributions of individuals included in this study.

|  | Indicated | Spontaneous | All Preterm | Term | Overall |
| --- | --- | --- | --- | --- | --- |
|  | (N=418) | (N=449) | (N=973) | (N=9671) | (N=10643) |
| **Race** |  |  |  |  |  |
| 1:Single race-White | 171 (40.9%) | 205 (45.7%) | 426 (43.8%) | 4715 (48.8%) | 5141 (48.3%) |
| 2:Single race-Black | 55 (13.2%) | 44 (9.8%) | 108 (11.1%) | 523 (5.4%) | 631 (5.9%) |
| 3:Single race-Latina | 57 (13.6%) | 46 (10.2%) | 111 (11.4%) | 679 (7.0%) | 790 (7.4%) |
| 4:Single race-Asian/Pacific Islander | 79 (18.9%) | 100 (22.3%) | 200 (20.6%) | 2466 (25.5%) | 2666 (25.0%) |
| 5:Single race-Native |  | 1 (0.2%) | 1 (0.1%) | 13 (0.1%) | 14 (0.1%) |
| 6:Mul race-Latina + other race | 23 (5.5%) | 17 (3.8%) | 48 (4.9%) | 433 (4.5%) | 481 (4.5%) |
| 7:Mul race-other races | 10 (2.4%) | 11 (2.4%) | 23 (2.4%) | 253 (2.6%) | 276 (2.6%) |
| 8:Other race | 17 (4.1%) | 16 (3.6%) | 36 (3.7%) | 426 (4.4%) | 462 (4.3%) |
| 9:Unknown | 6 (1.4%) | 9 (2.0%) | 20 (2.1%) | 162 (1.7%) | 182 (1.7%) |
| **Maternal age** |  |  |  |  |  |
| Mean (SD) | 33.9 (6.1) | 34.0 (5.4) | 34.0 (5.64) | 34.5 (4.84) | 34.4 (4.92) |
| Median [Min, Max] | 35.0 [15.0,51.0] | 35.0 [14.0,55.0] | 35.0 [15.0, 54.0] | 35.0 [14.0, 55.0] | 35.0 [14.0, 55.0] |
| Missing | 0 (0%) | 0 (0%) | 0 (0%) | 1 (0.0%) | 1 (0.0%) |
| **Private Insurance** |  |  |  |  |  |
| 0:No | 54 (12.9%) | 40 (8.9%) | 102 (10.5%) | 579 (6.0%) | 681 (6.4%) |
| 1:Yes | 363 (86.8%) | 407 (90.6%) | 865 (88.9%) | 9050 (93.6%) | 9915 (93.2%) |
| unknown | 1 (0.2%) | 2 (0.4%) | 6 (0.6%) | 41 (0.4%) | 47 (0.4%) |
| **Maternal Education** |  |  |  |  |  |
| <12 years | 13 (3.1%) | 11 (2.4%) | 25 (2.6%) | 71 (0.7%) | 96 (0.9%) |
| 12 years | 124 (29.7%) | 76 (16.9%) | 218 (22.4%) | 1348 (13.9%) | 1566 (14.7%) |
| >12 years | 230 (55.0%) | 308 (68.6%) | 589 (60.5%) | 7550 (78.1%) | 8139 (76.5%) |
| unknown | 51 (12.2%) | 54 (12.0%) | 141 (14.5%) | 702 (7.3%) | 843 (7.9%) |
| **Diagnosis Count** |  |  |  |  |  |
| Number of unique diagnoses<br>Mean (SD) | 10.9 (13.4) | 8.8 (10.1) | 9.6 (11.8) | 7.3 (8.5) | 7.5 (8.9) |
| Median [Min, Max] | 6.0 [1.0,101.0] | 5.0 [1.0,88.0] | 5.0 [1.0,101.0] | 5.0 [1.0,118.0] | 5.0 [1.0,118.0] |
| **Diagnosis Time** |  |  |  |  |  |
| Medical diagnosis visit time before conception<br>Mean (SD) | 1.8 (1.6) | 1.7 (1.6) | 1.7 (1.6) | 1.5 (1.6) | 1.6 (1.6) |
| Median [Min, Max] | 1.3 [0.0,8.7] | 1.1 [0.0,9.0] | 1.2 [0.0,9.0] | 1.0 [0.0,21.7] | 1.0 [0.0,21.7] |
| Medical diagnosis first visit time before conception<br>Mean (SD) | 2.4 (1.9) | 2.1 (1.9) | 2.3 (1.9) | 1.9 (1.8) | 2.0 (1.9) |
| Median [Min, Max] | 2.0 [0.0,8.7] | 1.5 [0.0,9.0] | 1.7 [0.0,9.0] | 1.3 [0.0,21.7] | 1.4 [0.0,21.7] |

For each individual, we identified all phenotypes present in their EHR before conception (**Figure 1b**). We harmonized phenotypes into phecodes, a curated grouping of ICD codes intended to capture clinically meaningful concepts (**Figure S1**). We identified 1322 unique phecodes across the cohort, and individuals had an average of 8 unique phecodes in their record prior to conception (**Figure S2b)**. Most diagnoses (86.8%) were rare — occurring in fewer than 1% of patients (**Figure S3a**). Most medical visits with a diagnosis occurred within 2 years before conception (**Figure S2b**); over 95% of individuals’ EHR start date was less than 2.5 years before conception (**Figure S2c**), and the maximum EHR length was 21.7 years before conception **(Figure S3b)**.

### Diverse pre-conception phenotypes associate with PTB risk

We tested each of the 1322 phecodes present in the cohort for association with preterm vs. term birth using logistic regression with maternal age, maternal education, insurance status, and race as covariates (**Figure 1c**). Of the covariates, maternal education had the highest rate of missingness at 7.9%. Race was missing for 1.7% of the cohort, and insurance classification was missing for 0.4% of the cohort (Table 1). Missing values for maternal education, race, and insurance classification were encoded as a separate categorical variable. We adjusted for multiple testing by controlling the false discovery rate (FDR) at 5% using the Benjamini-Hochberg procedure. Finally, for each significant association we evaluated its robustness by removing a random individual with the diagnosis and recomputing the association (Methods and **Figure 1c**). We repeated this process 50 times. An association which was significant in every iteration is considered robust.

We identified 34 significant and robust preterm birth associations among the 1322 diagnoses tested in the logistic regression (**Figure 2**). As expected, the most significant associations were well-established risk factors: type 1 diabetes (adjusted *P* = 1.7 × 10^-14^, *OR* = 8 (95% CI 4, 12)), essential hypertension (adjusted *P* = 2.1 × 10^−12^, *OR* = 3.3 (95% CI 2.5, 4.5)), and type 2 diabetes (adjusted *P* = 8.1 × 10^−12^, *OR* = 4.6 (95% CI 3.1, 6.8)). Since the PTB cohort includes both spontaneous and indicated preterm deliveries, these associations likely reflect medical practice as well as potential intrinsic risk.

**Figure 2:**
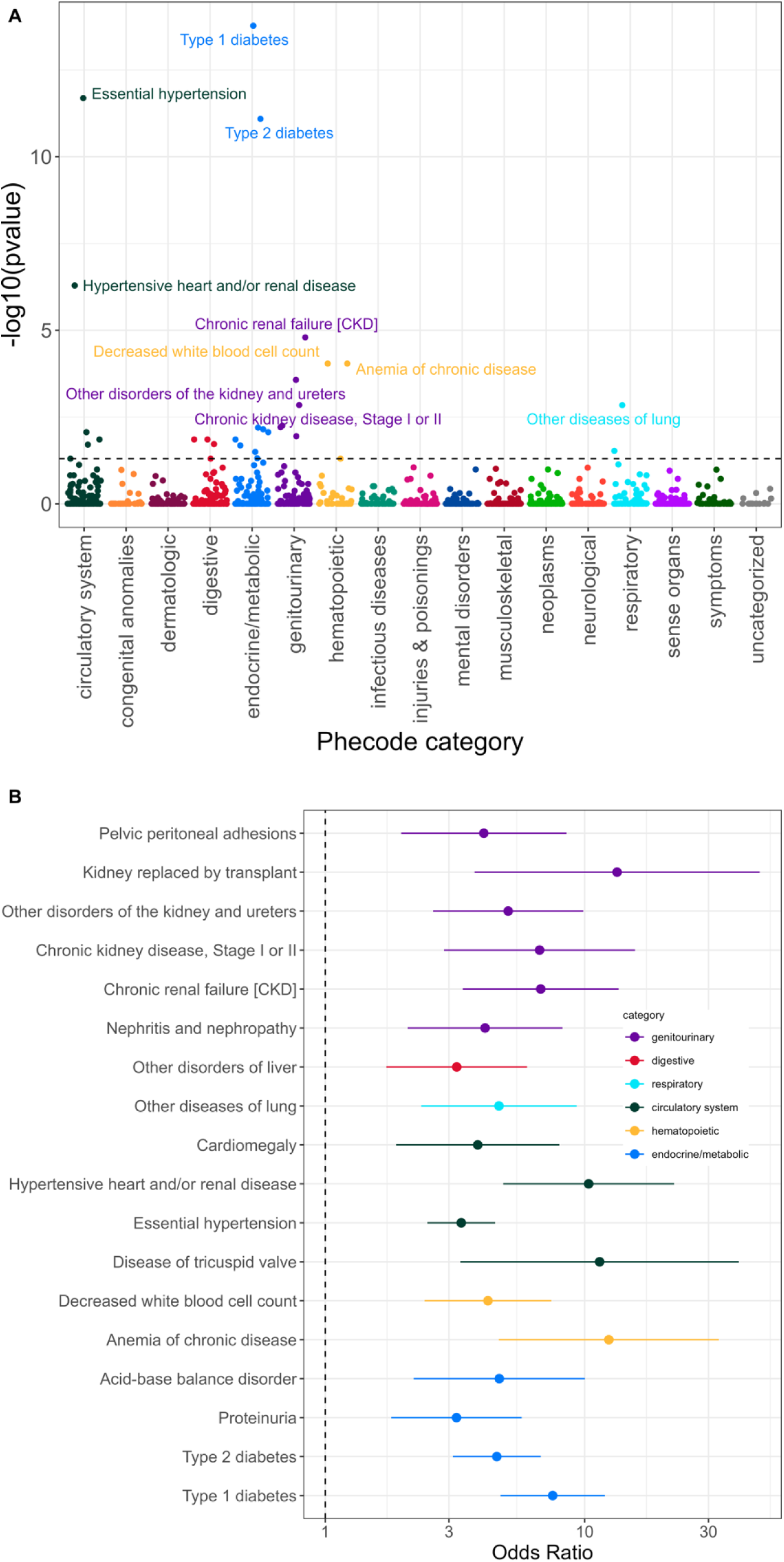
Testing Associations Between Clinical Phenotypes and PTB Identifies Known Risk Factors and Novel Candidates. **(A)** P-values from logistic regression tests of the association of 1322 clinical phenotypes with preterm (n=973) vs. term births (n=9671). Seventeen phenotypes passed the Benjamini Hochberg multiple testing corrected false discovery rate threshold of 5% (dashed line) and were robust to small changes in the data set. Phenotypes were represented as phecodes and plotted by phecode category. Significant, robust associations are labelled. **(B)** Forest plot shows odds ratios and 95% confidence intervals of the 17 conditions that were significantly and robustly associated with overall PTB. “Other disorders of liver” represents conditions including liver lesion, liver cirrhosis, liver mass, liver carcinoma, and fatty liver. “Other diseases of lung” represents conditions including lung consolidation, interstitial lung diseases, and lung mass.

After diabetes and hypertension related diagnoses, chronic kidney disease (CKD) was the next strongest preterm birth association (adjusted P = 1.6 × 10^−5^). Several other renal conditions were also among the significant associations, including a kidney replaced by transplant and other disorders of the kidney and ureters (**Figure 2b**). Associations between preterm birth and CKD have been observed in studies around the world, but the mechanisms and relevance to risk are not well understood (17).

The remainder of the significant associations included blood disorders, cardiac conditions, pulmonary conditions, liver conditions, electrolyte imbalances, and digestive conditions. To explore the meaning of the unspecific phenotypes “Other disorders of liver” and “Other diseases of lung,” we extracted concepts from clinical notes using ctakes (18). The “Other disorders of liver” (n=55) phenotype represents conditions including liver lesion (n=20), liver cirrhosis (n=15), liver mass (n=15), liver carcinoma (n=14), and fatty liver (n=13). The “Other diseases of lung” (n=38) phenotype represents conditions including lung consolidation (n=11), interstitial lung diseases (n=5), and lung mass (n=4). Odds ratios, P values, and sample sizes for associations between PTB and all 1322 diagnosis phenotypes are in **Figure 2** and **Table S1**.

### No diagnoses are associated with spontaneous preterm birth

Given the strong associations with preterm birth overall, we next tested for associations between clinical phenotypes and spontaneous preterm birth vs. term and indicated preterm birth vs. term. No diagnoses were significantly associated with spontaneous preterm birth (**Figure 3a**). In contrast, 30 diagnoses significantly and robustly associated with indicated PTBs (**Figure 3b**). The absence of significant associations with spontaneous preterm birth is not due to lower statistical power than for indicated preterm birth given their similar sample size (*nspontaneous* = 449, *nindicated* = 418).

**Figure 3:**
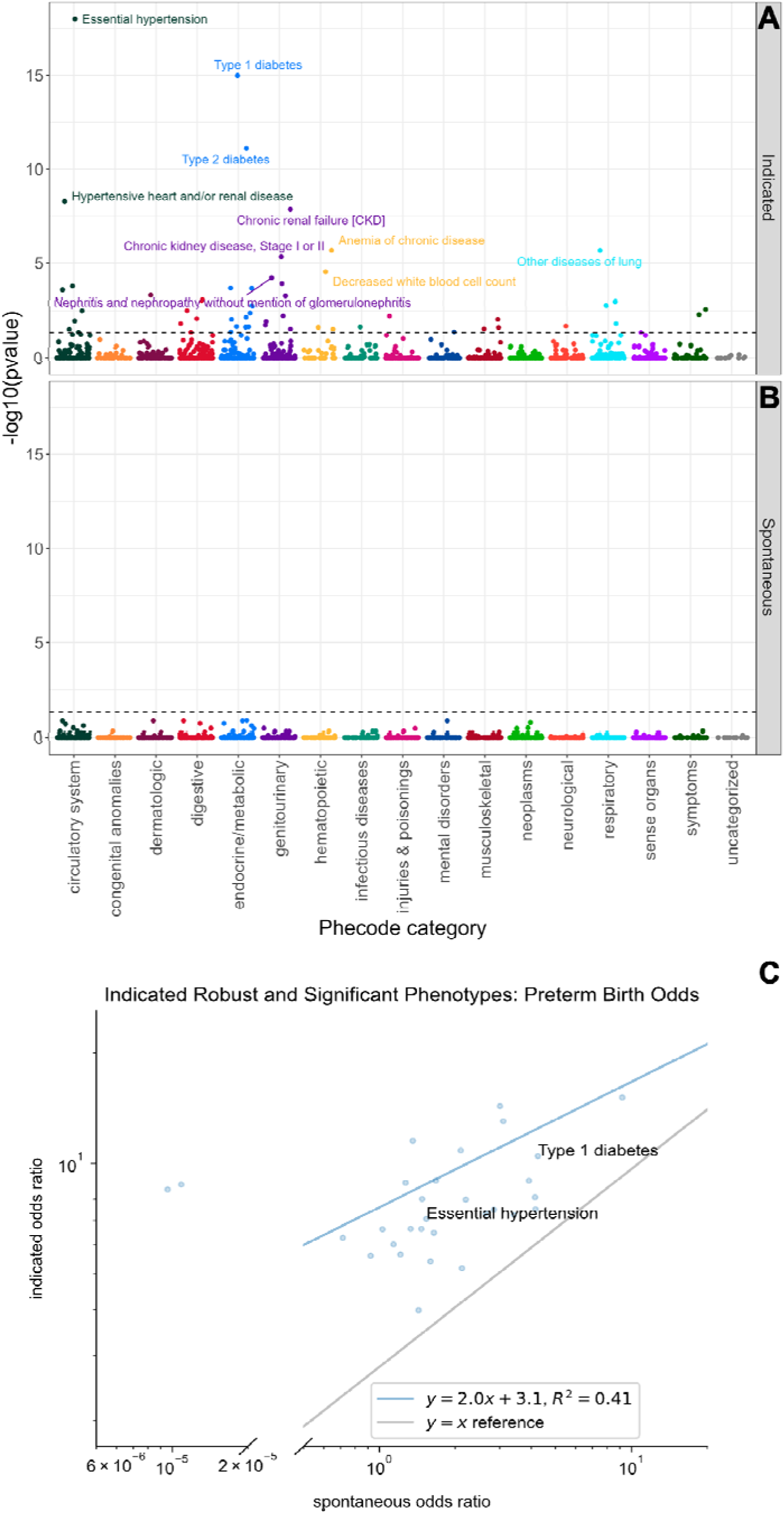
Many conditions associate with risk for indicated preterm birth, but none with spontaneous preterm birth. (A) P-values from logistic regression tests of the association of 1322 clinical phenotypes with indicated preterm (n=418) vs. term births (n=9671). Thirty phenotypes passed the multiple testing corrected Benjamini Hochberg threshold of 5% (dashed line) and were robust to small changes in the data set. (B) P-values from logistic regression tests of the association of 1322 clinical phenotypes with spontaneous preterm (n=449) vs. term births (n=9671). No phenotypes significantly associated with spontaneous preterm birth. (C) Comparison of the odds ratios for the 30 phenotypes significantly associated with indicated preterm birth between tests for indicated and spontaneous preterm birth. The odds ratios are correlated (r^2^=0.42, linear regression, left outliers dropped), but the relationships have systematically lower magnitude in the spontaneous cohort. The two most significant indicated phenotypes are labeled.

Of the 30 diagnoses associated with indicated preterm birth, 28 follow similar trends in spontaneous preterm birth, albeit at much lower effect sizes (r^2^=0.41, linear regression, **Figure 3c**). For example, hypertension has an odds ratio of 6 for indicated and 1.5 for spontaneous. The only two diagnoses with different directions of effect are acute laryngitis and tracheitis and congestive heart failure (CHF) not otherwise specified (NOS); both are significant, robust risk factors for indicated preterm birth, but are protective (though not significant) for spontaneous preterm birth.

### Phenotypes associated with all PTBs are also found for indicated PTBs

Of 18 significant and robust diagnoses associated with all PTBs, 17 are also among the 30 significant diagnoses associated with indicated PTBs (**Figure 4**). Diabetes, kidney diseases, and hypertension were the main diagnosis categories associated with overall and indicated PTB. The diagnoses associated with only indicated PTB but not the overall PTB cohort were spread across organs including the liver, lung, and heart. “Disease of tricuspid valve” was the only significant and robust association with overall PTB that was not found for indicated PTB (*PBH_indicated* = 0.28 *ORindicated* = 7 [1.2-42], *PBH_overall_preterm* = 8 × 10^−3^, *ORoverall_preterm* = 11 [3-39], *logistic regression*).

**Figure 4:**
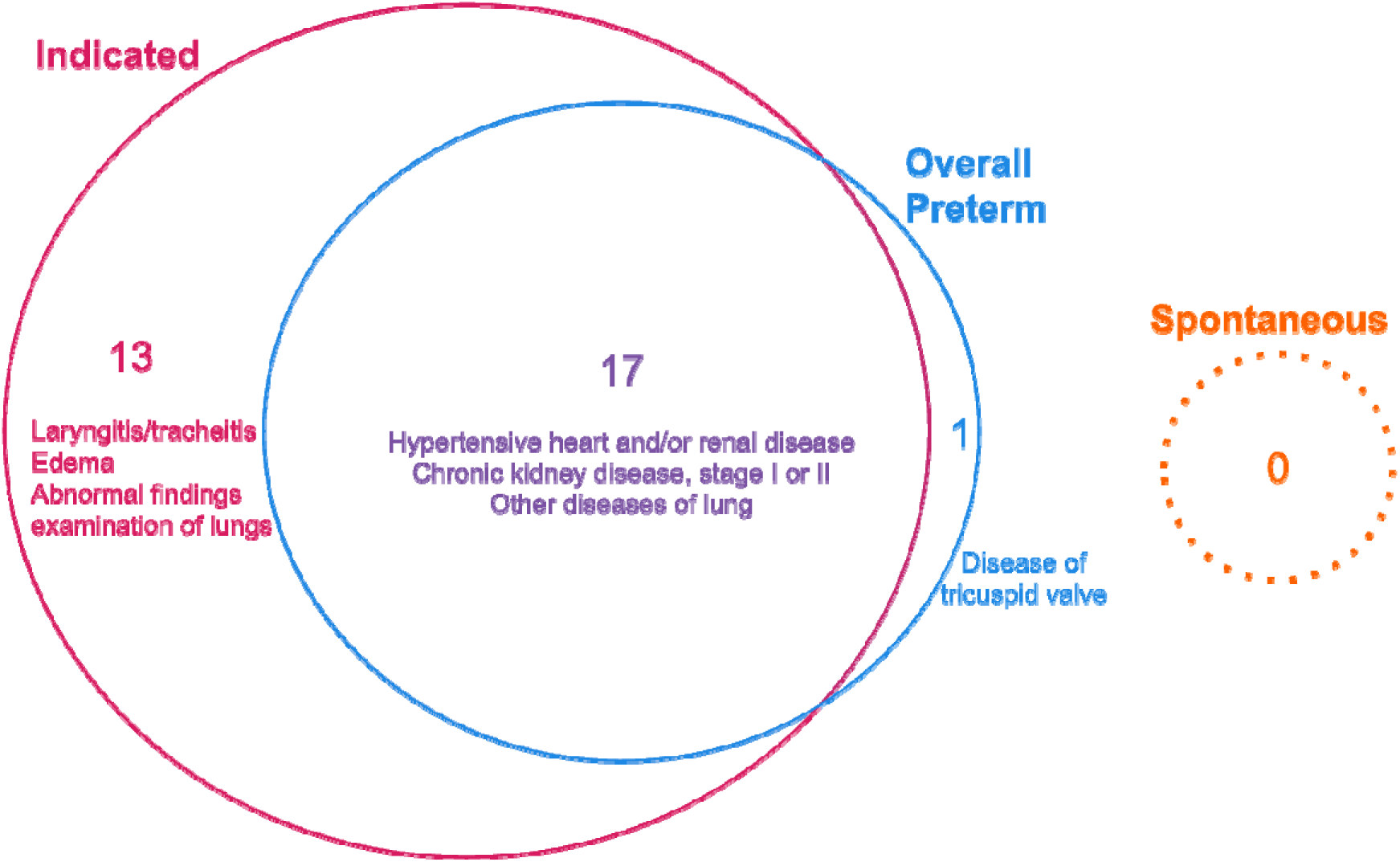
The strongest associations with overall preterm birth are also associated with indicated preterm birth, but not spontaneous preterm birth. Many kidney, cardiac, liver, and pulmonary conditions and diabetes are associated with overall and/or indicated preterm birth. Disease of tricuspid valve is a strong risk factor in overall preterm birth but not as strong in indicated preterm birth.

## 3 Discussion

Our study uses the rich phenotype data present in EHRs to generate hypotheses about the connection between diagnoses prior to pregnancy and risk for indicated and spontaneous PTB. We replicated known associations, including hypertension, diabetes, and chronic kidney disease. We also found several associations that warrant further investigation.

Most importantly, in our stratified analysis, we found no significant risk factors for spontaneous PTB. This underscores the limitations of approaches that combine indicated and spontaneous births together. When combined, significant associations were entirely driven by indicated PTBs, even though the spontaneous and indicated groups were of a similar size. Thus, our understanding of risk factors for spontaneous PTB remains limited.

The most significant hits of our study replicated well-established risk factors for PTB, with the four most significant being type 1 diabetes, essential hypertension, type 2 diabetes, and hypertensive heart and/or renal disease. These likely reflect clinical practice as they have existing recommendations for preterm delivery (9). Additionally, several significant phecodes relate to kidney function, such as chronic kidney disease, chronic renal failure, and other disorders of the kidney and ureters. Harel et. al propose that pre-pregnancy counseling, increased monitoring of the mother and fetus, and aspirin treatment to prevent preeclampsia would likely improve pregnancy outcomes for mothers with CKD, as indications for delivery are often hypertensive disorders, worsening renal function, fetal growth restriction or abnormal antenatal testing, or worsening maternal morbidity (17).

The association between decreased white blood cell count and overall PTB or related conditions is not widely agreed upon. Some studies found no association (19) while others did (20,21). The association between PTB and pre-conception lung conditions including lung consolidation, interstitial lung diseases, and lung mass is not well studied. Our association between PTB and pre-conception lung conditions coincides with the more established correlations that preterm birth causes lung conditions in the newborn through adulthood (22) and that preterm birth tends to pass down in families (23). Furthermore, the association between PTB and liver conditions including liver lesion, liver cirrhosis, liver mass, liver carcinoma, and fatty liver has been previously explored (24–26).

Comparing the results of the spontaneous subgroup analysis with the indicated subgroup analysis, we see that the two produce very distinct results. Further, we see that the indicated subgroup produces results very similar to the main analysis. This pattern is likely explained by the fact that established risk factors are key to clinicians’ decision-making when it comes to indicating delivery. As a result, we expect that the indicated group will have higher rates of these known risk factors and be more homogeneous. Our study highlights the fact that hypothesis-generation studies investigating PTB using data sources unable to distinguish between spontaneous and indicated PTB will have shortcomings. Investigating the heterogeneous spontaneous population has the potential to uncover lesser-known associations that may help reveal new pathways and understanding of PTB.

A major strength of our study is the use of a curated births database. This database not only records whether deliveries were spontaneous or iatrogenic (information which is not present in our EHR data), but also helps avoid outcome misclassification. Prior work has identified outcome misclassification as a concern for such EHR-based association studies (27). Additionally, researchers investigating obstetric data quality in an EHR system found that quality was varied and recommended manual abstraction where possible (28). By using a database reviewed by physicians to define our outcome, we minimize the risk of such misclassification.

An additional strength of our study is the wide range of phenotypes we were able to investigate. Our study covered more than 1,300 phenotypes across 17 different phecode categories.

There are a number of limitations in our study. Our data come from a tertiary care center, which presents two limitations: the patients and deliveries seen at this facility are not representative of the overall local population, and many patients being seen for delivery do not have a previous clinical record at the facility. We present both patient demographics (**Table 1**) and sample selection (**Figure 1**) to show the context in which the study was performed. Compared to the state of California, the PTB rate among our cohort is similar; however, our study population was generally older, more educated, and was privately insured at a high rate. The absence of a significant difference in age distribution between our indicated, spontaneous, and term cohorts may not reproduce in a US age-representative dataset (**Figure S2 a**). We do not have complete medical histories for the individuals included in our study; conditions or deliveries that occurred elsewhere may not be noted in our records.

Major chronic health conditions appear with high sample sizes and strong statistical power in our cohort. While we captured diagnosis phenotypes across all medical specialties using EHR, diagnosis phenotypes that are not major chronic health conditions will likely be underdiagnosed and under recorded. Consequently, we expected and found a lower sample size and weaker statistical power in our cohort for these phenotypes. More specifically, there are over 585 diagnoses that occur in fewer than 10 individuals (**Figure S3 a**), and over 300 of these are n_preterm_=0.

We expected that most diagnoses would be recorded in the short time before conception, and we found that over 50% of diagnoses occurred within 1 year of conception (**Figure S3 b**). This represents a common limitation of electronic health record trajectory analysis research, especially prevalent at UCSF as a tertiary care center: patients often use many healthcare institutions over their lifetime, and a patient’s medical history at any individual institution is usually missing data from previous institutions.

Our study is also limited by the generality of some diagnosis codes and insufficient disease quantification. For example, it is difficult to extract meaning from a phenotype named “Abnormal findings examination of lungs” that is too general and may represent undiagnosed diseases. Additionally, diagnoses do not indicate the severity of disease; a diagnosis of type 2 diabetes applies to both well-managed and poorly managed disease. However, in the poorly managed case, we are likely to see diagnoses representing additional complications. Future studies that supplement diagnosis binary data with severity data could provide insights about the relationships between disease severity and PTB. In EHR, this could be addressed with lab, vitals, and/or clinical notes data.

We propose two main areas of further research resulting from this work. The first is investigating lesser-known or unknown associations from our overall preterm analysis. Our work suggests several hypotheses which warrant more detailed study, especially in EHR systems complemented by other data sets. For instance, further work investigating the gastrointestinal associations would benefit from a combined EHR and gut microbiome biobank. The second area for future work is thorough investigation into the spontaneous group. We found no associations in our spontaneous subgroup, suggesting that this group may benefit from alternative approaches, such as dimensionality reduction and clustering, which may identify subtypes in the heterogeneity. We are also eager to study events and exposures during these pregnancies.

In a large EHR system, we replicated known associations between diagnosed conditions and PTB, as well as finding associations of interest for further study. We found that some pre-conception diagnoses were strong predictors of indicated PTB subgroup while all pre-conception diagnoses were poor at predicting spontaneous PTB. We hypothesize that there are strong associations between some diagnoses during pregnancy (e.g. life-threatening ones such as sepsis) and spontaneous PTB. These associations would likely be particularly prominent in our cohort of tertiary care individuals.

In this study, we only investigated one stratification of PTB: spontaneous/indicated. We ran preliminary studies on stratifying early (<32 weeks gestational age) and late (32-36 weeks) PTB, but the sample size for the early preterm group (n=132) was prohibitively small to produce any meaningful results on potential diagnoses associations. Future work should explore associations for different stratifications such as early/late preterm, young/mid/old maternal age, rural/suburban/urban maternal home, and low/middle/high maternal socioeconomic status. Identifying different risk factors and pathways to PTB could lead to better understanding of this heterogenous condition and to targeted and effective prevention efforts.

## 4. Methods

### Data Selection

#### Birth Data

We identified births using a perinatal database (PDB), which is maintained and curated by obstetricians at UCSF. This database contains detailed information about each delivery that takes place in the hospital and includes whether the delivery was spontaneous or indicated. Newborn patient IDs in this database are linked to newborn patient IDs in the EHR. The start of pregnancy was determined by subtracting gestational weeks from the delivery date.

#### Diagnosis Data

Diagnosis information was obtained from UCSF’s Observational Medical Outcomes Partnership (OMOP) de-identified EHR database, using mapped newborn patient IDs from the PDB. To be considered, diagnoses must have an ICD-9 or ICD-10 code and map to a phecode, and have a start date prior to the start of pregnancy. Conditions were considered to either be present or absent so that chronic conditions that may be recorded multiple times per pregnancy would not overwhelm our results. Phecodes were truncated after the first decimal point to provide an appropriate level of detail to conditions **(Figure S1)**.

#### Selection Criteria

We selected our sample from all deliveries at UCSF between 2001 and 2022. To be included, PDB maternal patient IDs must map to OMOP maternal patient IDs, and only one record per delivery may be present (**Figure 1a**). Additionally, deliveries must be singleton, have a recorded gestational age, have a recorded delivery date, and be from a birthing person with at least one diagnosis prior to the start of pregnancy.

#### Assigning conditions to pregnancies

For our main analysis, we included multiple deliveries from the same birthing person. Conditions were assigned per pregnancy in the following way: the condition must be diagnosed prior to the pregnancy start but not in the six-month period following the most recent delivery (**Figure 1b**). If no prior delivery was recorded, then conditions any time prior to the pregnancy were included.

#### Spontaneous and Indicated Preterm Definitions

A team of clinicians manually marked all 10,668 pregnancies in this cohort with a PTB status of “No” (this indicates a term birth), “spontaneous,” “PPROM,” “medically indicated,” “Termination Iatrogenic,” or “PTL with TOCO and TERM.” Additionally, pregnancies had data on gestational age, maternal age, maternal education level in years, and insurance type. Using the gestational age data, 975 of the newborns were delivered at fewer than 37 weeks, and 9,693 newborns were delivered at 37 weeks or more (**Figure 1a**). Pregnancies that were labeled with a gestational age of less than 37 weeks and a PTB status of “medically indicated” or “Termination Iatrogenic” were classified as indicated for this study. Pregnancies that were labeled with a gestational age of less than 37 weeks and a PTB status of “spontaneous,” “PPROM,” or “PTL with TOCO and TERM” were classified as spontaneous for this study.

In some cases, the PTB status value did not align with the gestational age value. In these cases, the pregnancies were dropped. Pregnancies marked with a gestational age of 37+ weeks and a spontaneous PTB status (n=18) represent individuals who experienced medically interrupted spontaneous preterm labor and delivered after term. It is unknown why some pregnancies would be marked with a gestational age less than 37 weeks and “No” PTB status (n=106).

#### Diagnosis-PTB Association Analysis

For all diagnoses occurring in at least one recorded birth, we implemented crude and adjusted logistic regression to test the associations between each diagnosis and unstratified PTB, indicated PTB, and spontaneous PTB.

#### Logistic Regression

Odds ratios and p-values were calculated by comparing spontaneous preterm pregnancies to term controls and indicated preterm pregnancies to term controls using logistic regression. We used the glm() function in R.

#### Covariates

Based on previous findings regarding PTB, we wanted to adjust for maternal age, socioeconomic status (SES), and racism. While we have maternal age as a variable, we can only use proxies for the latter two areas. For SES, we tested insurance status and maternal education. For racism, we used self-reported race and ethnicity. Our final covariates were maternal age, insurance (public vs private), education, and self-reported race and ethnicity. A smoothing spline was applied to maternal age to capture the non-linear relationship between age and PTB (29). Maternal education was reduced from the raw number of years value to the categories less than 12^th^ grade, 12^th^ grade, or college. As missingness for covariates was present in the data, unknown was considered to be a separate category for each.

#### P-Value Significance, Bootstrapping and Plotting

When classifying a phenotype as significant or not significant, p-values were adjusted for multiple hypothesis testing using the Benjamini Hochberg correction and tested against the threshold p_BH_<0.05 for 100% of 50 bootstrap iterations. Significant phenotypes with zero and infinity errors in the logistic regression (e.g. Intestinal infection, OR=1/∞, p=0, nindicated_preterm = 0, nterm = 30) were dropped.

As most of the diagnoses occur in fewer than 3% or 250 patients, it is important to consider that a small sample of individuals with a rare diagnosis could test as a significant association by chance instead of a causal relationship. Bootstrapping was performed on all analyses for 50 iterations of removing one instance of each diagnosis. For each iteration, a unique individual was selected. If the number of instances of the diagnosis was less than 50, then each instance was removed in exactly one iteration. If the number of instances of the diagnosis was 50 or greater, then each instance was removed at most in one iteration.

The Manhattan and Forest plots were generated using the R packages ggplot2 (version 3.4.2) (30) and ggrepel (version 0.9.3) (31). The odds-odds and supplementary figures were plotted using the Python packages Matplotlib (version 3.7.0) (32) and Seaborn (version 0.12.0) (33). Plots show adjusted p-values. Odds ratios and 95% confidence intervals are not adjusted for multiple hypothesis testing.

### 5 List of Abbreviations

EHR: Electronic Health Record
ICD: International Classification of Diseases
PTB: Preterm Birth
SES: Socioeconomic status
UCSF: University of California, San Francisco

## 6 Declarations

### Ethics Approval and consent to participate

This study was approved by the Institutional Review Board of University of California San Francisco (#17-22929).

### Consent for Publication

Not Applicable

### Availability of data and materials

Overall preterm, indicated preterm, and spontaneous preterm diagnosis association results are in Supplementary Files 1-3, respectively. In our association analyses, some diagnoses had very low (<10) patient counts. To maintain patient de-identification, exact counts, odds ratios, and p-values are redacted for those diagnoses in Supplementary Files 1-3. UCSF-affiliated individuals can request access to UCSF EHR data by contacting UCSF Information Commons.

The custom code/software we generated are available in the repository “stratified_PTB_association_study” available here [https://github.com/hanmochturt/stratified_PTB_association_study] This contains instructions for OMOP EHR data queries and all of the code for patient filtering, diagnosis aggregation, overall PTB association analysis, indicated and spontaneous PTB association analysis, healthcare time trajectory analysis, robustness testing, and figure creation.

## Funding

We would like to acknowledge the T32 institutional training grant (5 T32DE007306) and March of Dimes for funding.

### Authors’ Contributions

Conceptualization, JMC, JR, AT, TO, JAC, and MS; Methodology, JMC, HT, JR, AT, TO, JAC, and MS; UCSF EHR data access, JMC; Formal Analysis, JMC and HT; Clinical Insights, OY; Writing – Original Draft, JMC and HT; Writing – Review & Editing, JMC, HT, JAC, and MS; Visualization, JMC and HT; all authors read and approved the final manuscript.

## Data Availability

Overall preterm, indicated preterm, and spontaneous preterm diagnosis association results are in Supplementary Files 1-3, respectively. To maintain patient de-identification, exact counts, odds ratios, and p-values are redacted for diagnoses affecting >10 people in Supplementary Files 1-3.
The custom code/software we generated are available in the repository stratified_PTB_association_study available here [https://github.com/hanmochturt/stratified_PTB_association_study] This contains instructions for OMOP EHR data queries and all of the code for patient filtering, diagnosis aggregation, overall PTB association analysis, indicated and spontaneous PTB association analysis, healthcare time trajectory analysis, robustness testing, and figure creation.

## Acknowledgements

We would like to acknowledge Timothy Wen for his clinical insight, Melissa Rosenstein for access to the Perinatal Database, and the UCSF Information Commons team for EHR data access, members of the Sirota and Capra Labs for useful discussion.

## SUPPLEMENTAL FIGURES

### 7 Supplementary Information

**1.1 Supplemental Figures**

**Figure S1:**
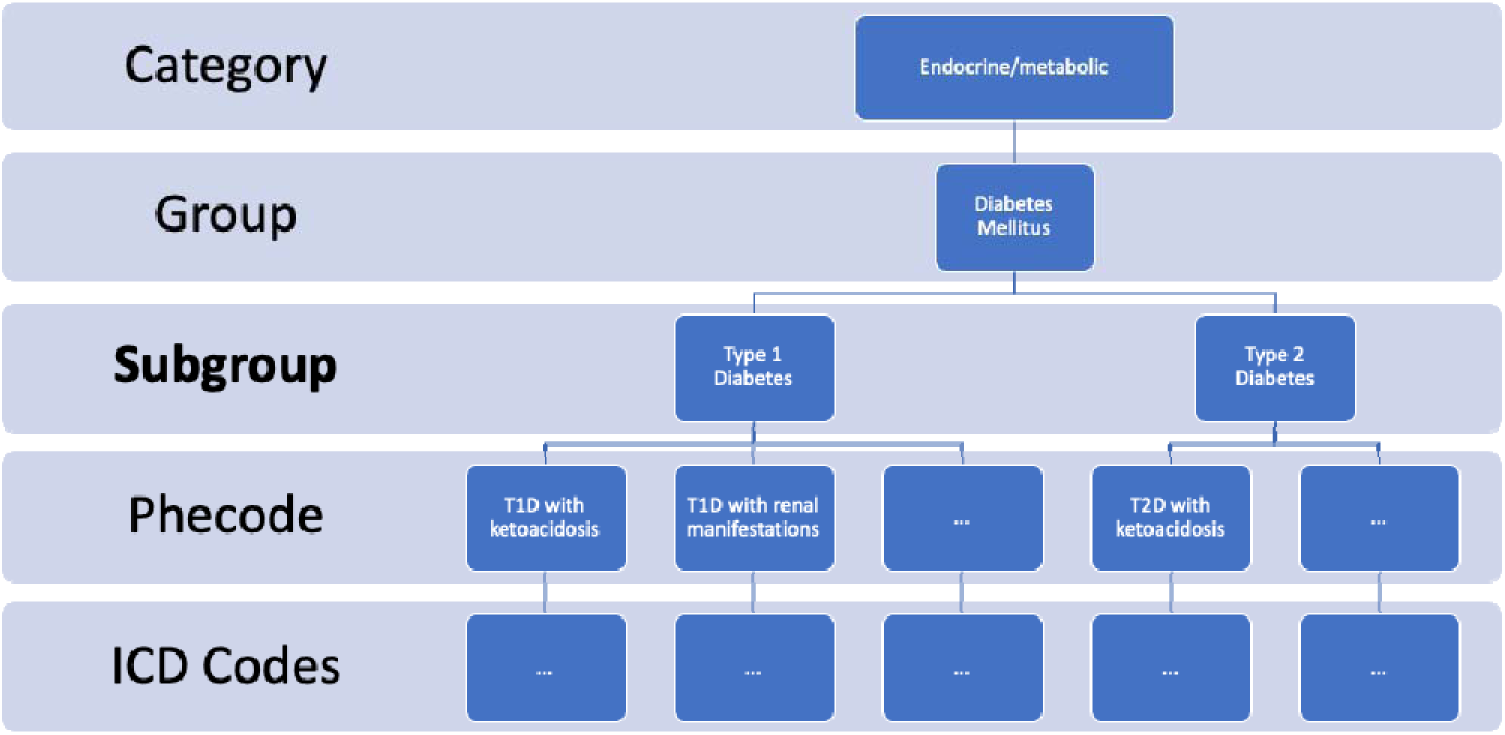
Phecode subgroups used to define diagnosis phenotypes. We use phecode subgroups to define diagnosis phenotypes. This allows us to test diagnosis-preterm associations with sufficient detail.

**Figure S2:**
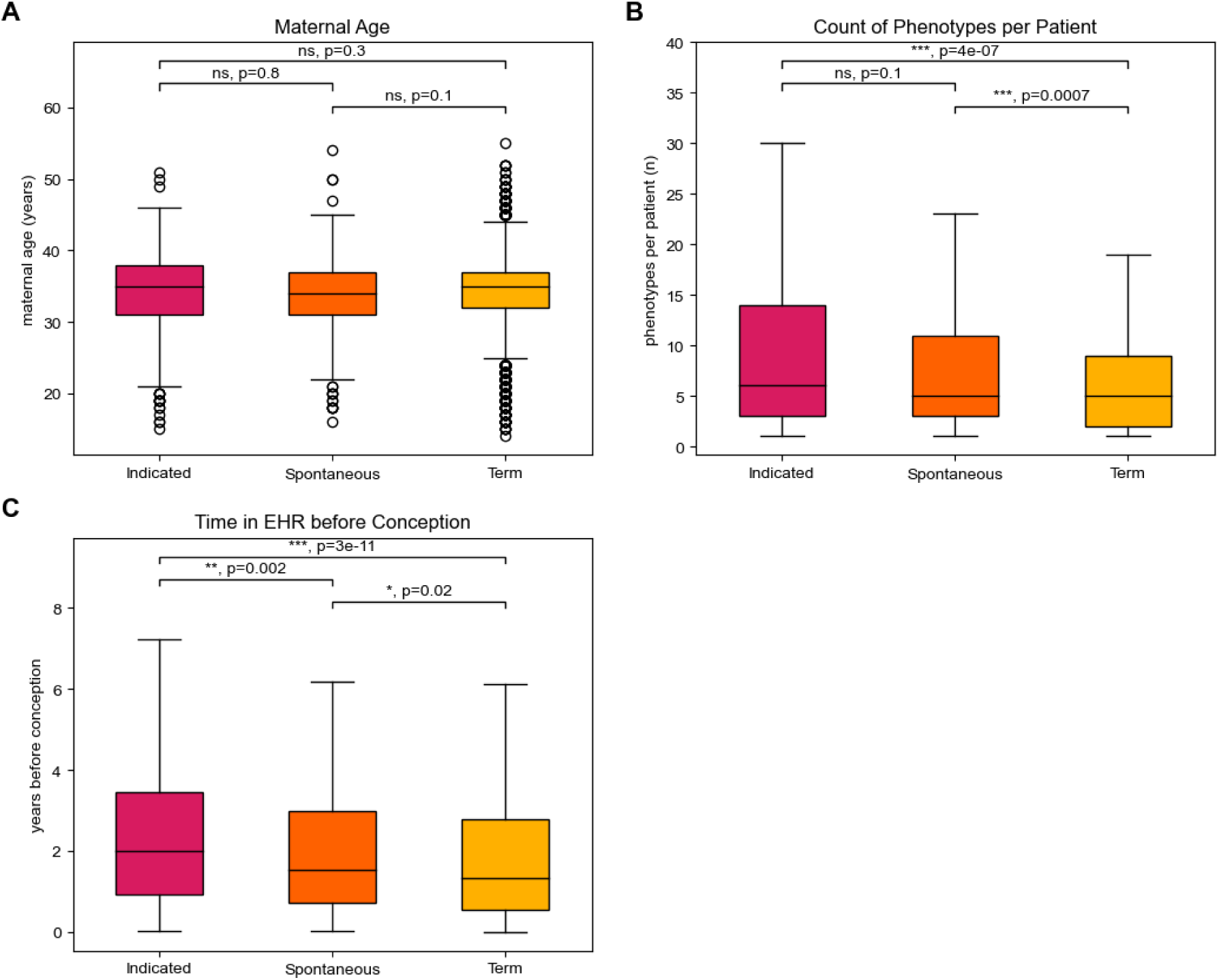
Patient Distributions for Maternal Age, Number of Diagnoses per Patient, First Visit per Patient, All Visits per Patient. **(Ai)** By 2-sided Mann-Whitney U rank test, there are no significant differences in maternal age between indicated, spontaneous, and term. **(B)** Additionally, by the same statistics test, outliers dropped, the spontaneous preterm and indicated preterm patients have significantly more diagnoses than the term patients. (C) Indicated individuals have the longest (time) EHR length, followed by spontaneous individuals, then term individuals with the lowest.

**Figure S3:**
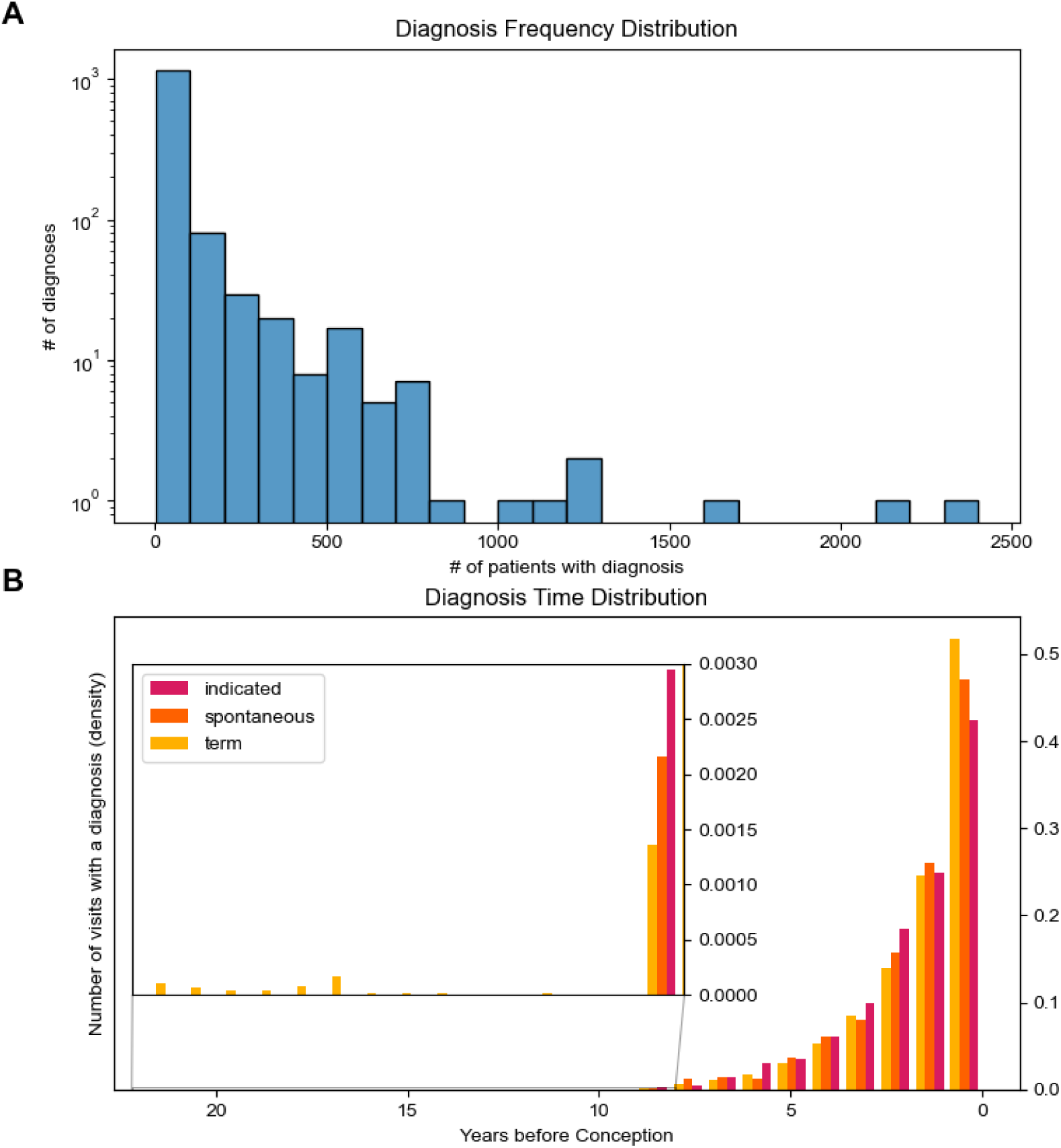
Diagnoses Frequency Distribution. **(A)** Most (1,148 of 1,322) diagnosis phenotypes occur rarely (in fewer than 100/10642 patients). **(B)** Most medical visits with a diagnosis occurred within 2 years before conception.

## References

1. Walani SR. Global burden of preterm birth. Vol. 150, International Journal of Gynecology and Obstetrics. 2020.

2. Behrman RE, Butler AS. Mortality and acute complications in preterm infants. Preterm birth: causes, consequences & prevention. 2006;

3. Moster D, Lie RT, Markestad T. Long-Term Medical and Social Consequences of Preterm Birth. New England Journal of Medicine. 2008;359(3).

4. Kramer MS, Lydon J, Goulet L, Kahn S, Dahhou M, Platt RW, et al. Maternal stress/distress, hormonal pathways and spontaneous preterm birth. Paediatr Perinat Epidemiol. 2013;27(3).

5. Andrews WW, Hauth JC, Goldenberg RL. Infection and preterm birth. Am J Perinatol. 2000;17(7).

6. Brou L, Almli LM, Pearce BD, Bhat G, Drobek CO, Fortunato S, et al. Dysregulated biomarkers induce distinct pathways in preterm birth. BJOG. 2012;119(4).

7. Jelliffe-Pawlowski LL, Baer RJ, Blumenfeld YJ, Ryckman KK, O’Brodovich HM, Gould JB, et al. Maternal characteristics and mid-pregnancy serum biomarkers as risk factors for subtypes of preterm birth. BJOG. 2015;122(11).

8. Goldenberg RL, Culhane JF, Iams JD, Romero R. Epidemiology and causes of preterm birth. Vol. 371, The Lancet. 2008.

9. Medically Indicated Late-Preterm and Early-Term Deliveries. ACOG Comm Opin. 2021 Jun 24;138(1).

10. Valencia CM, Mol BW, Jacobsson B, Simpson JL, Norman J, Grobman W, et al. FIGO good practice recommendations on modifiable causes of iatrogenic preterm birth. International Journal of Gynecology and Obstetrics. 2021;155(1).

11. Zhang YJ, Shen J, Lin SB, Lu C, Jiang H, Sun Y, et al. The risk factors of preterm birth: A multicentre case–control survey in China in 2018. J Paediatr Child Health. 2022;58(8).

12. Purisch SE, Gyamfi-Bannerman C. Epidemiology of preterm birth. Vol. 41, Seminars in Perinatology. 2017.

13. Malawa Z, Gaarde J, Spellen S. Racism as a root cause approach: A new framework. Pediatrics. 2021;147(1).

14. Kistka ZAF, Palomar L, Lee KA, Boslaugh SE, Wangler MF, Cole FS, et al. Racial disparity in the frequency of recurrence of preterm birth. Am J Obstet Gynecol. 2007;196(2).

15. Paquette AG, Hood L, Price ND, Sadovsky Y. Deep phenotyping during pregnancy for predictive and preventive medicine. Sci Transl Med. 2020;12(527).

16. Abraham A, Le B, Kosti I, Straub P, Velez-Edwards DR, Davis LK, et al. Dense phenotyping from electronic health records enables machine learning-based prediction of preterm birth. BMC Med. 2022 Dec 1;20(1).

17. Harel Z, Park AL, McArthur E, Hladunewich M, Dirk JS, Wald R, et al. Prepregnancy renal function and risk of preterm birth and related outcomes. CMAJ. 2020;192(30).

18. Savova GK, Masanz JJ, Ogren P V., Zheng J, Sohn S, Kipper-Schuler KC, et al. Mayo clinical Text Analysis and Knowledge Extraction System (cTAKES): Architecture, component evaluation and applications. Journal of the American Medical Informatics Association. 2010;17(5).

19. Musilova I, Pliskova L, Gerychova R, Janku P, Simetka O, Matlak P, et al. Maternal white blood cell count cannot identify the presence of microbial invasion of the amniotic cavity or intra-amniotic inflammation in women with preterm prelabor rupture of membranes. PLoS One. 2017;12(12).

20. Koleva-Korkelia I. MATERNAL LEUKOCYTOSIS AS DIAGNOSTIC MARKERS IN SPONTANEOUSLY DECLARED PRETERM BIRTH. Proceedings of CBU in Medicine and Pharmacy. 2021;2.

21. Bartkeviciene D, Pilypiene I, Drasutiene G, Bausyte R, Mauricas M, Silkunas M, et al. Leukocytosis as a prognostic marker in the development of fetal inflammatory response syndrome. Libyan Journal of Medicine. 2013;8(1).

22. Bolton CE, Bush A, Hurst JR, Kotecha S, McGarvey L. Lung consequences in adults born prematurely. Vol. 70, Thorax. BMJ Publishing Group; 2015. p. 574–80.

23. Koire A, Chu DM, Aagaard K. Family history is a predictor of current preterm birth. In: American Journal of Obstetrics and Gynecology MFM. 2021.

24. Zhuang X, Cui AM, Wang Q, Cheng XY, Shen Y, Cai WH, et al. Liver Dysfunction during Pregnancy and Its Association With Preterm Birth in China: A Prospective Cohort Study. EBioMedicine [Internet]. 2017 Dec 1;26:152–6. Available from: 10.1016/j.ebiom.2017.11.014

25. Amadou C, Nabi O, Serfaty L, Lacombe K, Boursier J, Mathurin P, et al. Association between birth weight, preterm birth, and nonalcoholic fatty liver disease in a community-based cohort. Hepatology [Internet]. 2022 Nov 1;76(5):1438–51. Available from: 10.1002/hep.32540

26. You S, Cui AM, Hashmi SF, Zhang X, Nadolny C, Chen Y, et al. Dysregulation of bile acids increases the risk for preterm birth in pregnant women. Nat Commun [Internet]. 2020;11(1):2111. Available from: 10.1038/s41467-020-15923-4

27. Beesley LJ, Mukherjee B. Statistical inference for association studies using electronic health records: handling both selection bias and outcome misclassification. Biometrics. 2022;78(1).

28. Altman MR, Colorafi K, Daratha KB. The Reliability of Electronic Health Record Data Used for Obstetrical Research. Appl Clin Inform. 2018;9(1).

29. Hutcheon JA, Moskosky S, Ananth C V., Basso O, Briss PA, Ferré CD, et al. Good practices for the design, analysis, and interpretation of observational studies on birth spacing and perinatal health outcomes. Paediatr Perinat Epidemiol. 2019;33(1).

30. Wickham H. ggplot2: Elegant Graphics for Data Analysis [Internet]. Springer-Verlag New York; 2016. Available from: https://ggplot2.tidyverse.org

31. Slowikowski K. ggrepel: Automatically Position Non-Overlapping Text Labels with “ggplot2” [Internet]. 2023. Available from: https://CRAN.R-project.org/package=ggrepel

32. Hunter JD. Matplotlib: A 2D graphics environment. Comput Sci Eng. 2007;9(3):90–5.

33. Waskom M. seaborn: statistical data visualization. J Open Source Softw. 2021 Apr 6; 6(60):3021.

